# Co-circulating pathogens of humans: A systematic review of mechanistic transmission models

**DOI:** 10.1101/2024.09.16.24313749

**Authors:** KE Shaw, JK Peterson, N Jalali, S Ratnavale, M Alkuzweny, C Barbera, A Costello, LT Emerick, GF España, AS Meyer, S Mowry, M Poterek, C de Souza Moreira, Eric Morgan, S Moore, TA Perkins

## Abstract

Historically, most mathematical models of infectious disease dynamics have focused on a single pathogen, despite the ubiquity of co-circulating pathogens in the real world. We conducted a systematic review of 311 published papers that included a mechanistic, population-level model of co-circulating human pathogens. We identified the types of pathogens represented in this literature, techniques used, and motivations for conducting these studies. We also created a complexity index to quantify the degree to which co-circulating pathogen models diverged from single-pathogen models. We found that the emergence of new pathogens, such as HIV and SARS-CoV-2, precipitated modeling activity of the emerging pathogen with established pathogens. Pathogen characteristics also tended to drive modeling activity; for example, HIV suppresses the immune response, eliciting interesting dynamics when it is modeled with other pathogens. The motivations driving these studies were varied but could be divided into two major categories: exploration of dynamics and evaluation of interventions. Finally, we found that model complexity quickly increases as additional pathogens are added. Future potential avenues of research we identified include investigating the effects of misdiagnosis of clinically similar co-circulating pathogens and characterizing the impacts of one pathogen on public health resources available to curtail the spread of other pathogens.

## 1. Introduction

Mathematical modelling is a powerful tool in understanding and forecasting infectious diseases and shaping public health responses (Ferguson et al. 2006, Heesterbeek et al. 2015, Pagel and Yates 2022). Mathematical models allow researchers to investigate the population-level impact of different treatment regimens for infectious diseases (Abu-Raddad et al. 2009), analyze the effect of nonpharmaceutical interventions on disease spread (Cowger et al. 2022), and predict how factors such as climate change will influence transmission of vector-borne diseases (Lafferty 2009, Eikenberry and Gumel 2018, Mordecai et al. 2019). However, the majority of infectious disease models focus on a single pathogen, despite the fact that multiple pathogens are co-circulating in any given time and place (Tsai et al. 2017, Mendenhall et al. 2022).

Co-circulating pathogens have the potential to impact the dynamics of one another via multiple mechanisms. People infected with HIV, for example, are more susceptible to infection by other pathogens, and this individual-level phenomenon can have population-level effects on transmission dynamics (Galvin and Cohen 2004, Mayer et al. 2007). HIV, TB, and malaria in particular co-circulate widely in under-resourced settings and can exacerbate both the clinical presentation and transmission of one another (Vitoria et al. 2009). This can lead to more interventions necessary at the individual-patient level, as well as a greater population-level burden of disease (Alemu et al. 2013, Montales et al. 2015). Furthermore, environmental factors such as climate change and land use changes likely impact multiple pathogens simultaneously and potentially synergistically, which could not be captured with a single-pathogen focus.

Deforestation and the expansion of agriculture, for example, have been shown to shift the abundance, diversity, and biting rates of mosquito species that are known arbovirus vectors (Zahouli et al. 2017, Da Silva Pessoa Vieira et al. 2022).

Shifting public health priorities in response to new threats can also have unintended effects on co-circulating pathogens. For example, during the height of the COVID-19 pandemic, routine vaccination campaigns across the world against diseases such as measles and polio were disrupted, with ripple effects on vaccine coverage and transmission dynamics (Gaythorpe et al. 2021, Cardoso Pinto et al. 2022, Bigouette et al. 2022). In addition, co-circulating pathogens with similar clinical presentations, such as Zika, dengue, and chikungunya, may hinder surveillance efforts to discern the true prevalence of each pathogen contributing to the observed syndromic data due to misdiagnosis and also hampering targeted public health interventions (Oliveira et al. 2020, Oidtman et al. 2021, Ribas Freitas et al. 2024).

Given the many ways in which co-circulating pathogens may impact each other, mathematical models that explicitly model these interactions may provide key insights. For example, researchers may be better able to capture transmission dynamics and thereby improve forecasting accuracy by including synergistic interactions such as one pathogen increasing host susceptibility to other pathogens (Marimuthu et al. 2020), or mitigating interactions such as infection causing decreases in host mobility, thus decreasing host exposure to other co-circulating pathogens (Rohani et al. 2003). The presence of co-circulating pathogens in a population may necessitate different screening or treatment regimens to achieve public health goals, and mathematical models can aide in exploring these different scenarios (Macgregor et al. 2020, Rao et al. 2022). The myriad possibilities in these approaches motivated us to explore the current state of the literature on mathematical models of co-circulating pathogens.

Despite the evidence for the impact of co-circulating pathogens on each other and the potential usefulness of mathematical models, we did not find any systematic reviews of mathematical models of co-circulating pathogens in our literature search. Therefore, we conducted this systematic review with the goal of characterizing the existing scientific literature on co-circulating pathogen models, to both better understand the current state of this field and identify knowledge gaps that need to be addressed. Specifically, our objectives were:

i. To determine the most common pathogens modeled together and the ecological, biological, and clinical similarities between these pathogens,
ii. To analyze the authors’ motivations for creating co-circulation models and the intended purpose of their studies,
iii. To characterize common features of co-circulation models and their outputs,
iv. To quantify differences between single-pathogen and co-circulating pathogen models, and
v. To report aspects of co-circulation models that are uncommon in the literature and potentially fruitful avenues for future research.

## 2. Methods

### 2.1 Literature search

On October 21^st^, 2022 we searched Google Scholar, Pubmed, and Web of Knowledge databases using a set of keywords. Our keyword list consisted of: *co-circulating pathogen; concurrent epidemic; polymicrobial infection AND mathematical model; concurrent infection AND mathematical model; syndemic AND model; co-infection AND mathematical model; heterologous reactivity AND mathematical model; polyparasitism AND mathematical model; synergistic pathogens AND mathematical model; dual infection AND mathematical model; pathogen co-occurrence; co-epidemic; superinfection AND mathematical model*. We retrieved a total of 1,908 unique bibliographic records from Google Scholar, PubMed, and Web of Science, which we reviewed in a four-stage process. In stage one, the titles and abstracts of the resulting manuscripts were briefly scanned for the inclusion criteria, which yielded a total of 343 papers published between 1985 and 2023 (1,565 excluded). In stage two, the references of each selected paper were then scanned to find additional publications, which yielded an additional 206 papers. Additionally, any newly published and relevant paper that the authors became aware of during the review process were added (five papers total). In stage three, a pair of reviewers assessed each paper (N=554) for meeting our inclusion criteria (below) and reached a consensus. In stage four, the reviewers first extracted the data separately and then later compared their questionnaire answers to reach a consensus on each question. We excluded papers not published in English (Figure 1).

**Figure 1.**
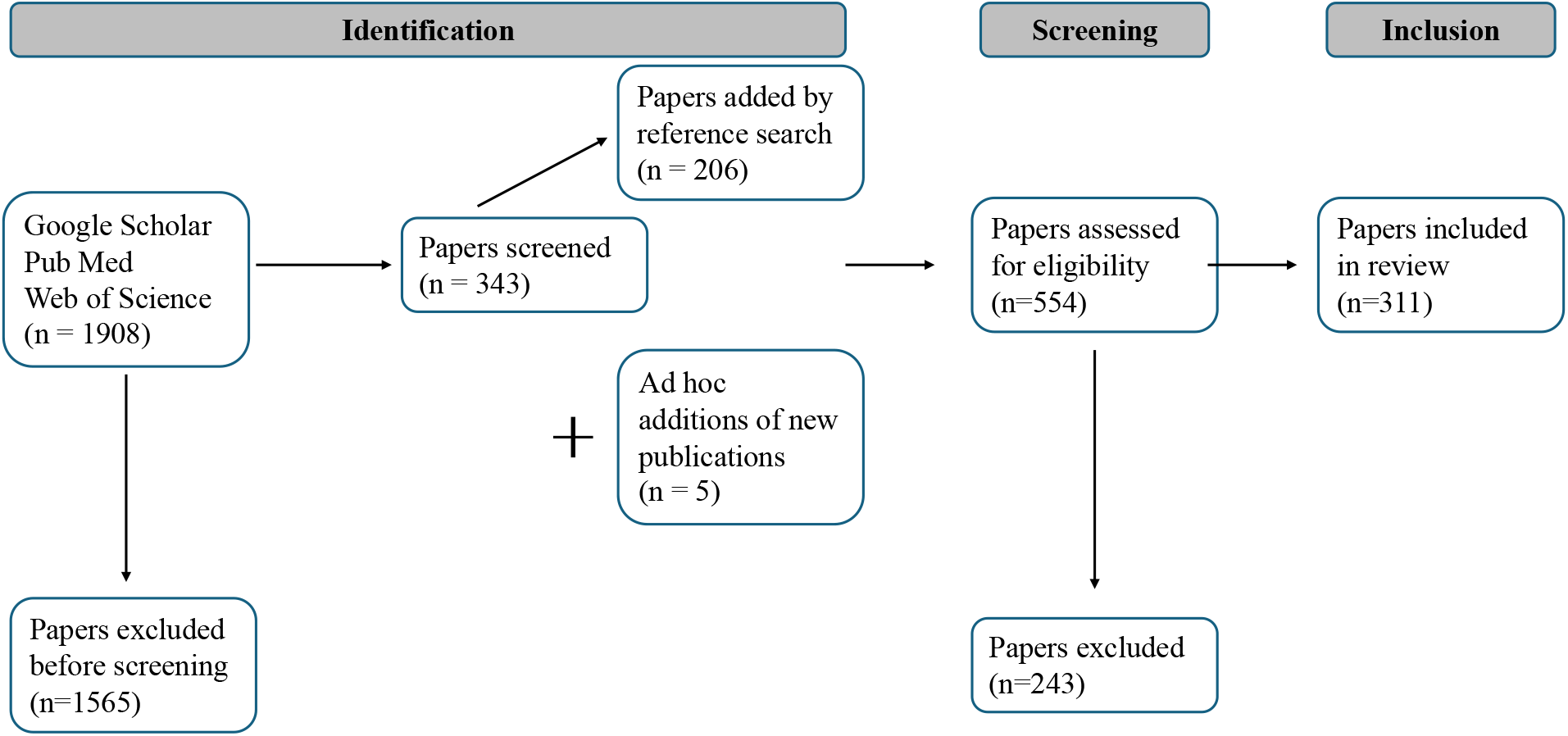
PRISMA flowchart for the literature search, screening, and inclusion process.

### 2.2 Eligibility criteria

All 554 papers were evaluated for inclusion based on six questions:

i. Is the paper an original research article?
ii. Are at least two real pathogen species (not theoretical) modeled in the same model?
iii. Does at least one of the pathogens modeled cause human disease?
iv. Does the model have at least one population-level aspect in a human population?
v. Is some part of the model structure informed by a mechanistic derivation of biological processes as opposed to purely involving phenomenological relationships?
vi. Is there a place in the model where hosts transition from a susceptible to an exposed or infected state?

We narrowed the scope of this review to real pathogen species that cause human disease (criteria ii and iii) because co-circulating pathogens have been a salient topic in the non-human literature for a longer period, and we were interested in how that has percolated into the modeling of human disease. We focused on mechanistic, population-level models that include a transmission process (criteria iv-vi) in contrast to descriptive or statistical models that fit data without any consideration of the biological mechanisms involved. We included papers for which the answer to all six questions was “yes”.

### 2.3 Questionnaire development

We used a questionnaire to screen each publication in order to standardize the data generated from each review. We developed the questionnaire through a collaborative, iterative three-stage process. In the first stage of the development process, our aim was to evaluate the ability of the questionnaire to elicit similar responses from multiple reviewers assessing the same paper. To this end, reviewers used the questionnaire to review the same paper, and we then evaluated response data for discrepancies. Through group discussion, we identified areas of improvement through group discussion, and feedback from the group was applied to the next iteration of the questionnaire. We carried out this process twice. Finally, in the third round of evaluation all papers were reviewed in pairs as described below and the data generated were used for analysis. All reviewers were in communication throughout the review stage to harmonize any discrepancies that arose.

### 2.4 Paper review process

Two reviewers were randomly assigned to each publication, which they reviewed using the standardized questionnaire. After reviewing each paper individually, the pair of reviewers assigned to each paper then met to compare and discuss answers and resolve any discrepancies between answers. Each pair then submitted a final version of their answers in the form of a third survey marked ‘consensus.’ Only consensus surveys were analyzed (N=311).

### 2.5 Questionnaire description

The questionnaire consisted of six sections:

i. bibliographic information;
ii. inclusion criteria;
iii. basic information about each parasite modeled;
iv. model details;
v. model outputs; and
vi. motivation for selecting the pathogens modeled. The final version of the questionnaire contained 47 questions.

### 2.6 Author motivations

Question 39 of our systematic survey was an open-response question: “Did the authors develop their model to answer a specific research question or to achieve a specific purpose or objective?” To analyze the responses to this question, we inserted the pooled responses to this question and into ChatGPT version 3.5 (“ChatGPT” 2024) with the prompt “The text I provided you was supposed to illustrate authors’ motivations to write the papers we analyzed. From that text, what would you say are the main motivations of authors?” Authors KS and TAP then discussed the output from ChatGPT and refined the language of the motivations ChatGPT produced to create four categories of motivations, and KS categorized each response to question 39 according to these motivations. Each response could receive more than one categorization.

### 2.7 Bibliometrics

To explore how journal prestige and topic may be related to citation patterns, for all papers included in our review for which a DOI was available, we queried Unpaywall (Richard Orr, Heather Piwowar, Jason Priem 2024) and OpenAlex (Aria et al. 2024) to collect the times each paper has been cited as well as the h-index and i-10index for the journal in which each paper was published, as of March 11^th^, 2024. We then used the glmmTMB package in R (Brooks et al. 2017) to create generalized linear models to assess potential associations between citation count and publication year, journal h-index, and journal i-10index. We also used the Web of Science API Lite (“Web of Science API” 2024) to collect subject categories for each paper. We then binned related categories such as “Mathematics, Applied” and “Mathematics” to create seven meta-categories. We created a second generalized linear model to test for the impact of assigned meta-category on citation counts.

### 2.8 Complexity Index

To estimate the extent to which each co-circulating pathogen model in our data set diverged from a single-pathogen model, we designed a complexity index derived from the responses to questions 23-30 in the questionnaire. The index was calculated by assigning one point to each answer that indicated a divergence from a single-pathogen model and then totaling the points.

For example, question 23 (“Does the model structure include hosts that are co-infected with two or more pathogens?”) would have yielded one point for a “yes” answer and zero points for a “no.” For multi-part questions such as question 23f (“Which parameter values for the co-infected class are different from the mono-infected and uninfected classes? [select all that apply]”), we assigned one point for each parameter value selected. The maximum possible complexity index score was 18. An example of how this index was calculated can be found in Table S1.

### 2.9 Data availability

A full version of the questionnaire can be found in the supplementary materials. All data were analyzed in R version 4.3.0 (R Core Team 2021), and raw files and code used for analysis as well as a supplementary file of all tables produced from the analysis are available at https://figshare.com/s/2b268f5df8aed6acad3f.

## 3. Results

### 3.1 Study selection

In our four-stage review process, we identified 554 papers to review for eligibility, 311 of which met our inclusion criteria (Figure 1). The most common reasons for paper exclusion were: (i) the study did not include two real pathogen species in the same model (67.1%, 163/243, question 7);(ii) there was no place in the model where hosts transitioned into an exposed or infected state (42.4%, 103/243, question 11); and (iii) the model did not have at least one population-level aspect (35.4%, 86/243, question 9). Some papers were excluded based on multiple criteria. The oldest papers that met our search criteria were from 1991, although the majority of relevant papers were published in 2008 or later (90.4%; 281/311, Figure 2).

**Figure 2.**
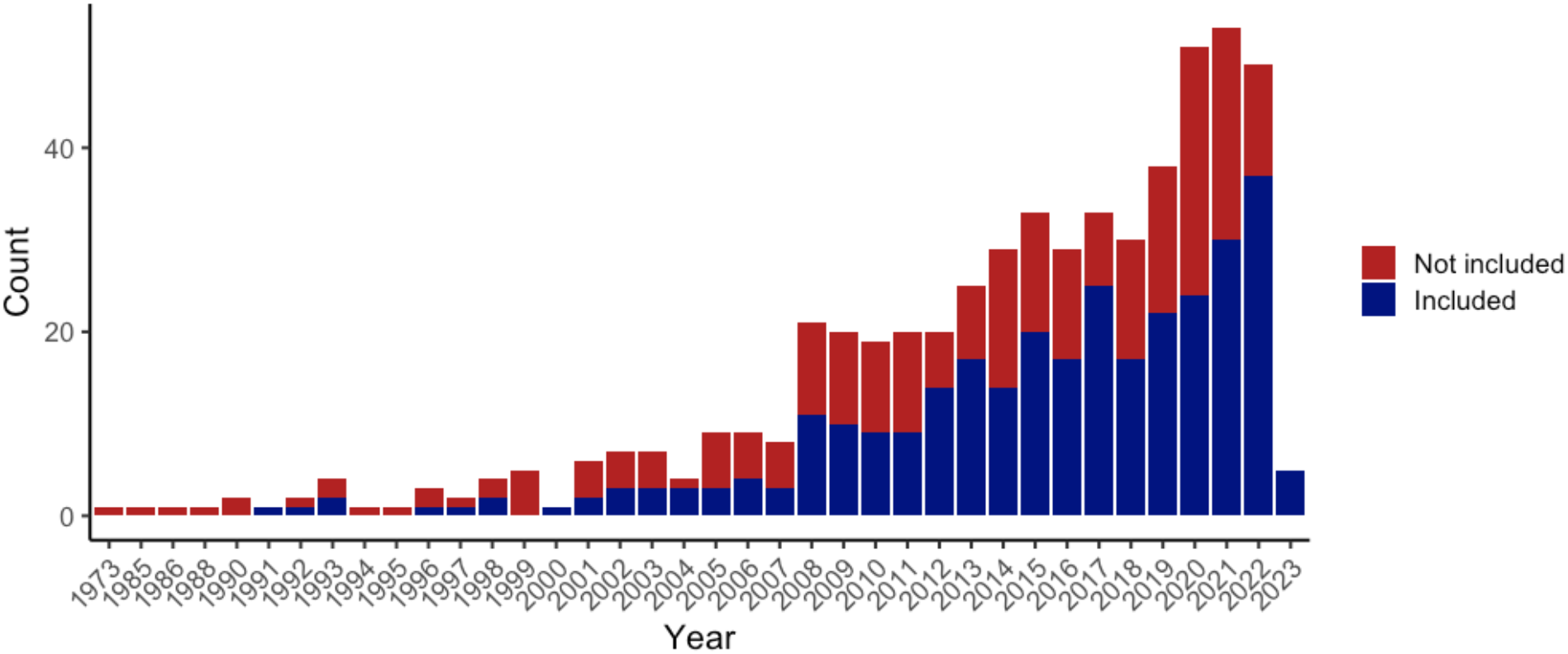
Temporal trends in the publication of co-circulating pathogen models. Papers that met our inclusion criteria gradually increased over time. The year of 2023 was only partially sampled up to papers published by the month of March.

### 3.2 Pathogen information

The majority of included models considered two pathogens (88.4%, 275/311), while the maximum number of pathogens included in the same model was nine (0.32%, 1/311; Figure 3A). The most commonly modeled pathogen pairs were human immunodeficiency virus (HIV) and *Mycobacterium tuberculosis* (TB), representing 25% (78/311) of the models we reviewed. Other models that included more than two pathogens often were focused either on STIs (e.g., Korenromp et al. 2005, Kate K Orroth et al. 2007, Jenness et al. 2017) or vector-borne diseases such as dengue, chikungunya, and Zika that are transmitted by the same vector (e.g., Isea and Lonngren 2016, Okuneye et al. 2017). One model that included nine pathogens considered a panoply of common STIs: HIV, genital herpes, syphilis, cancroid, gonorhea chlamydia, trichomonas, bacterial vaginosis, and vaginal candidiasis (Johnson et al. 2011).

**Figure 3.**
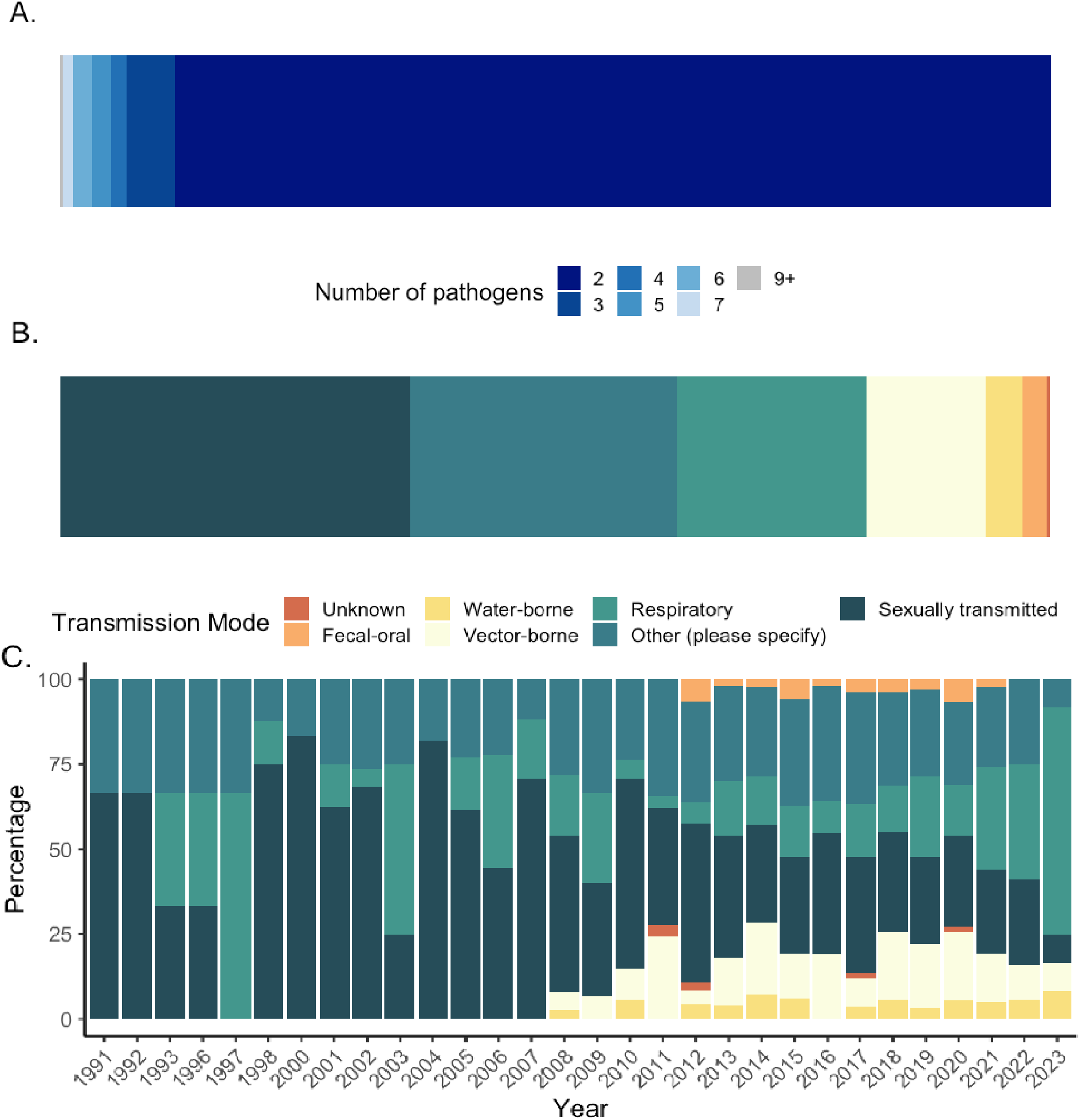
Pathogen characteristics. **(A)** The frequency of how many pathogens were included in models. The vast majority of models included two pathogens. **(B)** Frequency of transmission modes represented across all models. Sexually transmitted pathogens and pathogens with “other” transmission routes such as bloodborne were common in our data set due to the large number of models focused on HIV. **(C)** Change in transmission modes represented in our data set over time, represented as a percentage of the total models we analyzed for that publication year. Vector-borne pathogens appear more frequently in the literature after 2008, and respiratory pathogens become more common starting in 2021. **(B)** and **(C)** share a legend.

To investigate which modes of transmission were included in the same models, we recorded all viable transmission modes for each pathogen; e.g., HIV would be recorded as both “sexually transmitted” and “other (vertical and bloodborne).” Therefore, when we calculated the percentage of each transmission mode in our data, the denominator was the total number of transmission modes recorded across all pathogens (988). Corresponding to the exceptionally high number of HIV-TB models, the most common transmission modes of pathogens were sexually transmitted (35.3%, 349/988), followed by a transmission mode of “other” (27%, 267/988) of which 65.5% (175/267) were recorded as vertical and bloodborne (Figure 3B). Respiratory transmission was the third most common transmission mode at 19% (188/988). We also found that the earliest papers in our data set tended to focus on sexually transmitted and respiratory pathogens, again reflecting the focus on HIV in combination with other STIs and with TB in the 1990s (Figure 3C). Over time, a greater diversity of transmission modes appeared, notably with an increase in models of vector-borne diseases starting in 2008 and an increase in models with respiratory pathogens starting in 2020, coinciding with the SARS-CoV-2 pandemic.

Beyond individual pathogens, we also investigated the frequency with which different pathogen taxa were modeled together. We categorized the taxonomic group and transmission mode for each pair of pathogens in our data set and counted the frequency of their co-occurrence in the same model (Figure 4). This analysis reflected all possible transmission routes for each pathogen and also all possible pairings within the same model. As such, a single paper could be represented multiple times based on the number of pathogens and transmission modes present, resulting in 957 distinct pathogen pairs. Sexually transmitted viruses with sexually transmitted bacteria (11.4%, 109/957) and sexually transmitted viruses with viruses with other transmission routes (11.3%, 108/957) occurred together most commonly, due to the high prevalence of STI- focused models in our review. Other common pairings were respiratory bacteria with sexually transmitted viruses (9.1%, 87/957) and respiratory bacteria with viruses with other transmission routes (9.2%, 88/957), due to the large number of HIV-TB models.

**Figure 4.**
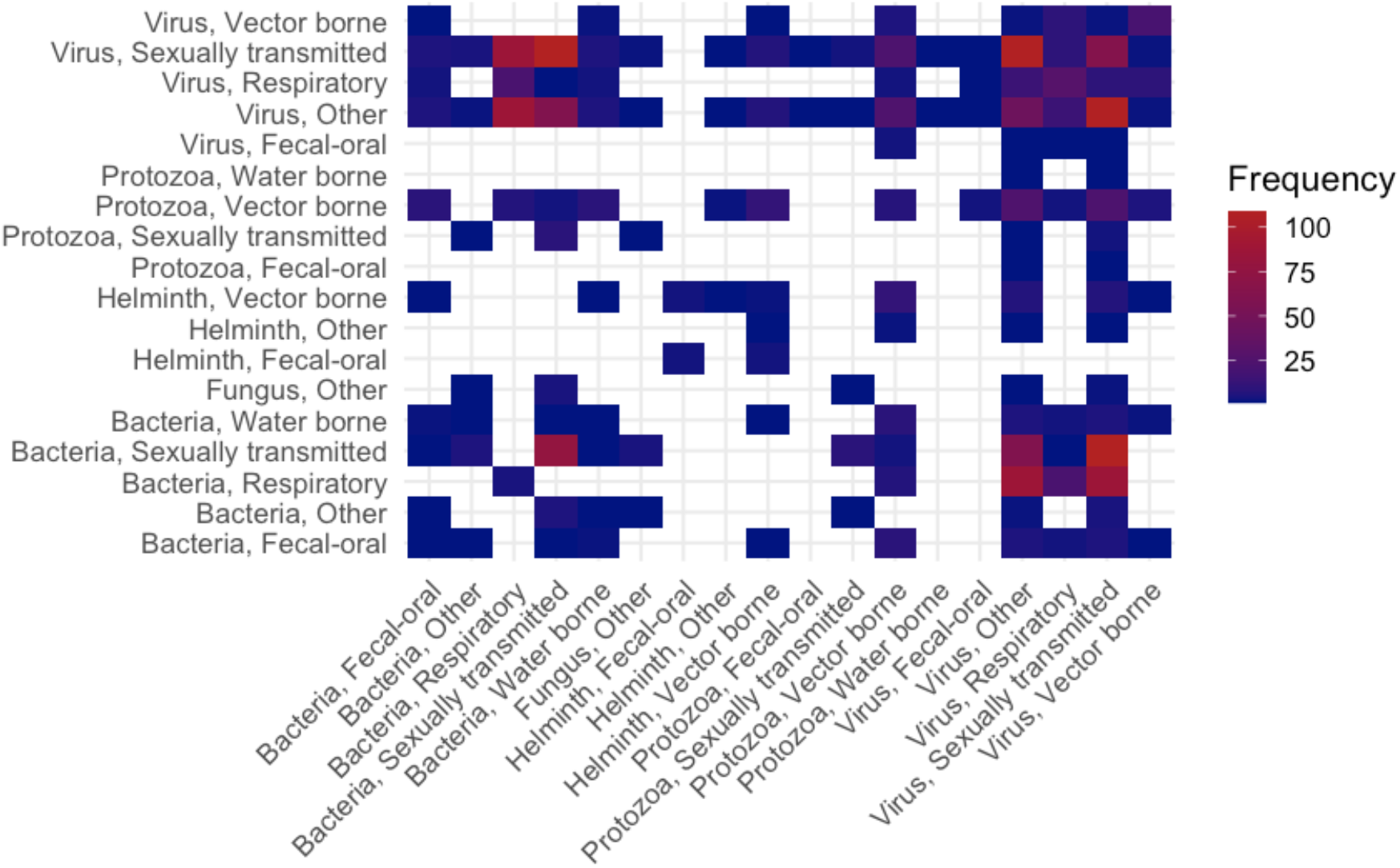
Frequency of co-modelling pathogens within taxonomic groups and transmission modes. The most frequent pairings were sexually transmitted viruses with sexually transmitted bacteria and viruses with other transmission routes, followed by sexually transmitted viruses with respiratory bacteria (dominated by the prevalence of HIV-TB models in our data set).

We found that the majority of pathogens modeled together did not share ecological similarities, as reported by authors (87.8%, 273/311, question 40). Of those that did, 63.2% (24/38) had shared seasonality and 39.5% (15/38) had shared environmental drivers of transmission. Shared seasonality was commonly noted for respiratory pathogens such as SARS-CoV-2, influenza, and RSV. Shared environmental drivers were noted for pathogens such as malaria and cholera, which are both sensitive to environmental conditions (e.g., rainfall) but via different mechanisms (Thomson et al. 2006, Lemaitre et al. 2019). Furthermore, pathogens that shared both seasonality and environmental drivers were noted for several models (9/38, 23.7%, Figure S1A). Multiple ecological similarities were most often recorded for models of vector-borne pathogens with a shared vector taxa, such as the dengue, chikungunya, and Zika viruses.

In contrast, a majority of pathogens modeled together did share at least one biological similarity (55.6%, 173/311, question 41), with 70.5% sharing a transmission route (122/173) and 63 % (109/173) sharing host subsets as the most common responses. Shared transmission routes were common in models with sexually-transmitted pathogens, vector-borne pathogens, and respiratory pathogens. Models with shared host subsets commonly included studies of intravenous drug users (de Vos et al. 2012, Vickerman et al. 2012, Zelenev et al. 2018) and childhood diseases (Rohani et al. 2003, Huang and Rohani 2006). Clinical or epidemiological similarities were shared by 62.7% (195/311, question 42) of pathogens modeled together, with the most common similarity being that treatments existed for both pathogens (59%, 115/195), due to large number of HIV-TB and STI models for treatable pathogens.

### 3.3 Motivations and purpose

Reviewers identified two main scholarly motivators (question 45): 1) prior research suggested an effect of one pathogen on another but more research is needed (54.7%, 170/311), and 2) few or no prior modeling studies conducted of these two pathogens together (35.1%, 109/311). No clear scholarly motivation was noted in 11.6% (36/311) of papers.

In addition, we used ChatGPT (version 3.5) (“ChatGPT” 2024) to summarize common themes in the reviewers’ answers to the open-ended question 39, “Did the authors develop their model to answer a specific research question or to achieve a specific purpose or objective?” (Box 1). We then refined the results from ChatGPT to better reflect the nuances we felt existed in our data set, resulting in four categories of author motivation. We then classified reviewer responses to question 39 according to the four motivation categories, allowing for more than one category to apply to each paper. The categories that resulted from this process were:

i. **Understanding the dynamics of co-circulating pathogens**, in particular how multiple pathogens may alter the transmission patterns and/or population outcomes of one another. This category applied to papers in which the authors’ goals included evaluating quantitative outcomes such as R0, disease burden, mortality, etc., and how outcomes differed in the presence of co-circulating pathogens.
ii. **Developing mathematical models** for specific pairs of pathogens of interest, often due to the perceived novelty of doing so by the authors. While all papers in this review included a mathematical model, this category was for those that had a primarily mathematical focus or in which the authors only stated motivation for developing the model was that the pathogens had not been previously modeled together.
iii. **Implementing interventions or control measures** for one pathogen and the resulting impact on it and/or other pathogens in the model. We placed papers in this category when the authors were investigating how interventions such as treatment, vaccination, or quarantining would impact co-circulating pathogens through either direct or indirect effects.
iv. **Calculating cost-effectiveness and optimal control** of different treatment and control strategies. While there was some overlap with (iii), we created this category to capture papers in which the authors were not necessarily investigating the impact of interventions on outcomes such as transmission or mortality, but in which they were primarily concerned with the best way to implement interventions given cost or labor constraints.

Category (i), understanding disease dynamics, was the most common motivation for creating the models, with 52.3% (164/311) of papers falling into this category. Implementation of interventions (iii) was the second most common, at 35.4% (110/311), followed by cost-effectiveness (iv, 14.9%, 46/311) and developing mathematical models (ii, 13.8%, 43/311).

Overlapping categorizations were common, in particular for categories (i) and (iii); Fig S2). Some papers fell into one or more of the four categories but also contained unique motivations and thus were counted in one or more of the four main categories and additionally as “other” (10%, 31/311). A small number, 1.6% (5/311), did not neatly fit into any of these categories and were categorized only as “other.” Examples of “other” author motivations include examining how one pathogen may exert indirect evolutionary pressures on another (Lloyd-Smith et al. 2008), or what fundamentally allows for the co-existence of two co-circulating pathogens (Amaku et al. 2011).

#### Box 1.

Large language learning model analysis of survey question 39, “Did the authors develop their model to answer a specific research question or to achieve a specific purpose or objective?”

We provided the reviewers’ answers to question 39 for all papers included in our review that met inclusion criteria and provided the prompt “The text I provided you was supposed to illustrate authors’ motivations to write the papers we analyzed. From that text, what would you say are the main motivations of authors?” The exact output created, prior to our modified version stated in the text, was as follows:

1. “Understanding Disease Dynamics: Authors aim to gain a deeper understanding of the dynamics of infectious diseases, including their transmission patterns, interactions between different pathogens, and the impact of interventions on disease spread.
2. Developing Mathematical Models: Many authors are motivated to develop mathematical models to simulate disease transmission, assess the effectiveness of intervention strategies, and predict the outcomes of various scenarios. These models allow researchers to study complex disease dynamics and inform public health decision-making.
3. Evaluating Intervention Strategies: There is a strong emphasis on evaluating the effectiveness and cost-effectiveness of intervention strategies, such as vaccination programs, treatment protocols, and preventive measures. Authors seek to identify optimal strategies for controlling disease spread and reducing disease burden in populations.”

### 3.4 Bibliometrics

We were able to source bibliometric data for 247 out of 311 of the papers in our analysis. The majority of these papers were cited fewer than 25 times (64.3%, 159/247, Figure 5). However, there were some distinct outliers, with the most highly cited paper having 231 citations (Table 1). Year of publication (z value = -7.721, p<0.001) and h-index (z value = 3.501, p=0.0005) of the journal in which a paper was published were both significantly associated with citations, but the effect sizes were minimal (central estimates of -0.0857 per year and 0.001653 per h-index unit, respectively), suggesting that outliers in citations likely reflect papers that were particularly useful or interesting to the field.

**Figure 5.**
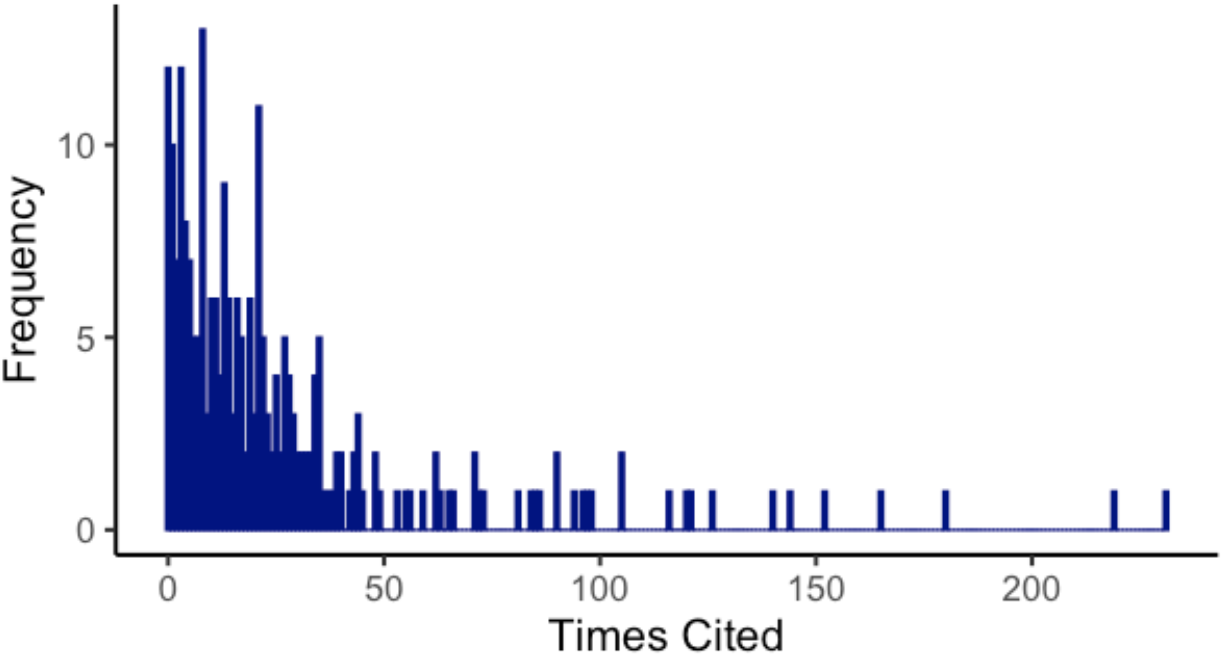
Frequency of citations. Most papers in our data set were cited fewer than 25 times, however there were several outliers with over 100 citations.

**Table 1.** Top 10 cited papers from our data set.

| Title | Journal | DOI | Times cited |
| --- | --- | --- | --- |
| Modelling the impact of global tuberculosis control strategies | *Proceedings of the National Academy of Sciences* | 10.1073/pnas.95.23.13881 | 231 |
| Genital herpes has played a more important role than any other sexually transmitted infection in driving HIV prevalence in Africa | *PLOS One* | 10.1371/journal.pone.0002230 | 219 |
| Incidence of Gonorrhea and Chlamydia following Human Immunodeficiency Virus preexposure prophylaxis among men who have sex with med: A modeling study | *Clinical Infectious Diseases* | 10.1093/cid/cix439 | 180 |
| Mathematical analysis of the transmission dynamics of HIV/TB coinfection in the presence of treatment | *Mathematical Biosciences and Engineering* | 10.3934/mbe.2008.5.145 | 165 |
| Ecological interference between fatal diseases | *Nature* | 10.1038/nature01542 | 152 |
| Determinants of the impact of sexually transmitted infection treatment on prevention of HIV infection: A synthesis of evidence from the Mwanza, Rakai, and Masaka Intervention Trials | *The Journal of Infectious Diseases* | 10.1086/425274 | 144 |
| Comparing Dengue and Chikungunya emergence and endemic transmission in *A. aegypti* and *A. albopictus* | *Journal of Theoretical Biology* | 10.1016/j.jtbi.2014.04.033 | 140 |
| Identifying the interaction between influenza and pneumococcal pneumonia using incidence data | *Science Translational Medicine* | 10.1126/scitranslmed.3005982 | 126 |
| Can Hepatitis C Virus (HCV) direct-acting antiviral treatment as prevention reverse the HCV epidemic among men who have sex with men in the United Kingdom? Epidemiological and modeling insights | *Clinical Infectious Diseases* | 10.1093/cid/ciw075 | 121 |
| Beneficial and perverse effects of isoniazid preventative therapy for latent tuberculosis infection in HIV-tuberculosis coinfecting populations | *Proceedings of the National Academy of Sciences* | 10.1073/pnas.0600349103 | 120 |

We found additional metadata for 244 of our 311 papers on the Web of Science search engine. This metadata provided subject categories for each paper, with one to five categorizations per paper. By binning related categories (for example, “Mathematics, Applied” and “Mathematics”; Table S2), we created seven main categories related to the research subject areas for each paper. Life sciences (54.1%, 132/311) and Mathematics (52.9%, 129/311) were the most common categories, while Operations and Management was the least common (3.3%, 8/311). Overlap between categories, in particular Mathematics and Life Sciences, was also common (Figure S3). Similar to citation year and journal h-index, we found that some categories were significantly associated with citations; again though, effect sizes (relative to Operations and Management) were minimal (Life Sciences: estimate = 0.639, z value = 3.207, p=0.00134; Medicine/Public health: estimate = 0.593, z value = 2.199, p=0.028; Multidisciplinary: estimate = 0.731, z value = 3.015, p = 0.00257).

### 3.5 Model Characteristics and Outputs

The majority of publications presented deterministic models (89.7%, 279/311), followed by individual-based models (8.4%, 26/311). A very small number of papers contained a within-host model in addition to the population-level model required to meet our inclusion criteria (3.9%, 12/311). Spatially explicit models and metapopulation models were rare among the papers reviewed (0.64%, 2/311 and 0.32%, 1/311, respectively). Some papers contained multiple model components with varied structures, and thus fell into multiple categories (6.4%, 20/311).

85.2% of the publications included models that were informed by data (265/311). Of these papers, 56.4% (149/265) used data extracted from the literature without giving further information on the origin of those data. Surveillance data were used in 29.1% (77/265) of papers, 10.2% (27/265) used survey data, and 3% (8/265) used case study data. The most common use of data was for model parameterization (97%, 257/265), including the use of data-informed parameter values taken from other modelling studies. A smaller set of papers used data for model fitting or calibration (36.2%, 96/265).

The most commonly reported model output (question 35) was disease prevalence (79.4%, 248/311), followed by the reproduction number (59.5%, 185/311) and stability properties of the model (57.9%, 180/311). Several model outputs were not initially captured by our survey options and were coded as “other” (49.2%, 153/311). Among these, common responses we found in the “other” category were model sensitivity analysis, cost evaluation of different interventions, and total cases of disease averted by interventions. Outputs were commonly examined as a function of different interventions (question 36, 46.6%, 145/311), time (22.5%, 70/311), and varying parameter values (22.5%, 70/311). Rarely did models evaluate outputs as a function of seasonality (0.64%, 2/311), space (0.32%, 1/311), or temperature/climate (0.32%, 1/311).

### 3.6 Complexity Index

We created a complexity index to quantify how much co-circulating pathogen models diverged from a set of independent, single-pathogen models. Scores were assigned based on questions 23- 30, with one point for each increase in complexity (see Methods). The largest score on the complexity index we observed was 11, out of a maximum possible value of 18. Scores in the intermediate range of 4-7 were most common (72.7%; 226/311), with very few at the minimum score of 0 (2/311) or the maximum observed value of 11 (2/311;Figure 6A). This middle range of complexity reflected a pattern we commonly observed where most models aimed to investigate one or two dimensions of co-circulation/co-infection, but upon the incorporation of a second (or more) pathogen(s), the biology of the interplay between the pathogens often required multiple changes to the model relative to a single-pathogen model. The most commonly observed elements of the complexity index were including hosts that were co-infected (94.5%, 294/311, question 23), including external factors that affect the transmission of both pathogens (e.g., interventions) (52.7%, 164/311, question 30), and allowing mono-infected hosts to have different levels of susceptibility to other pathogen(s) in the model as compared to uninfected hosts (52.4%, 163/311, question 24). The least commonly observed elements of the complexity index were accounting for interruptions or declines in services offered due to another co-circulating pathogen in the model structure (0.96%, 3/311, question 28) and the model structure including reporting or surveillance errors due to misdiagnosis between the pathogens modeled (1.3%, 4/311, question 26). Models that included within-host aspects of co-infection in the model structure were also rare (2.6%, 8/311, question 23a; Figure 6B).

**Figure 6.**
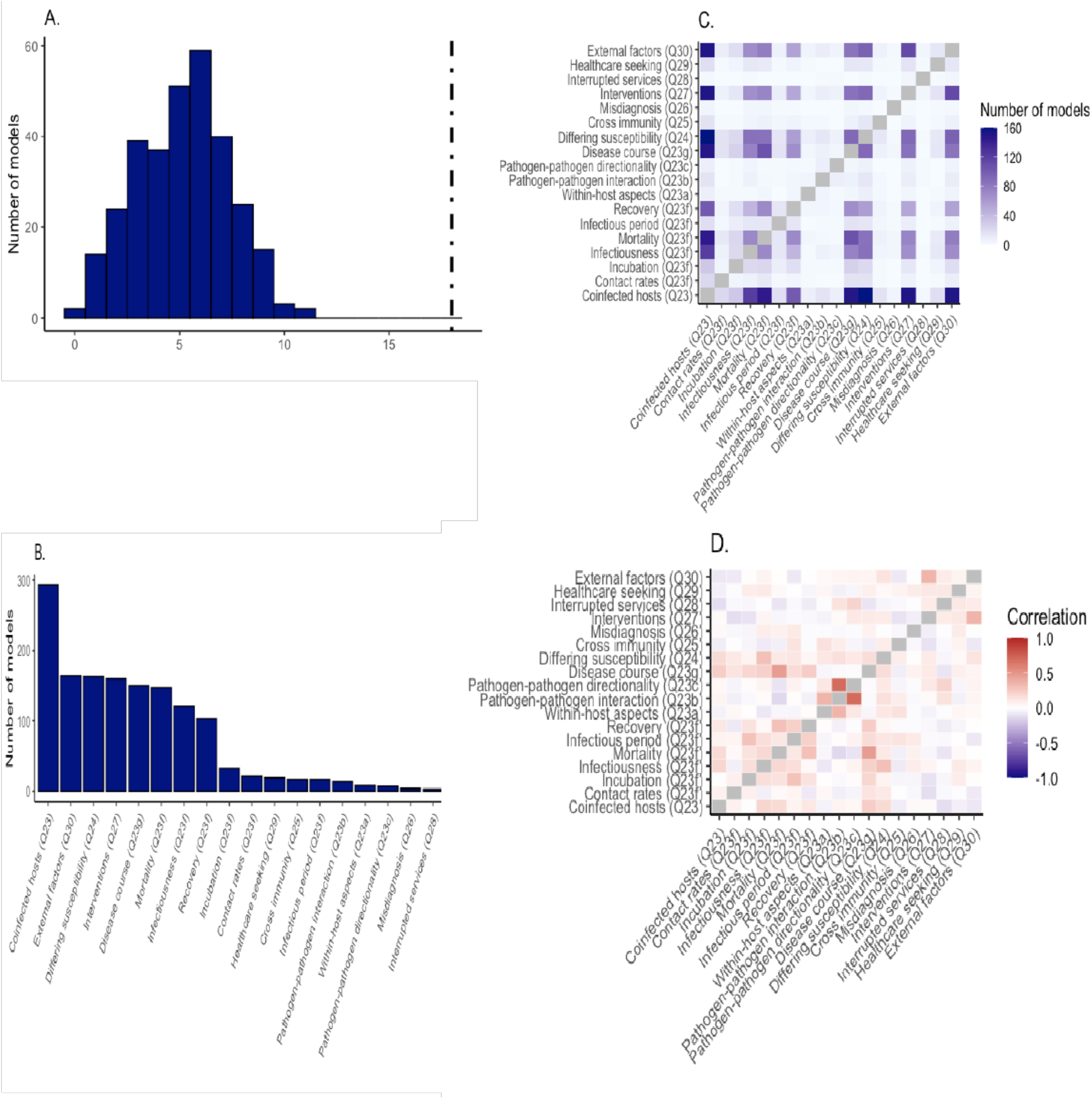
Measures of model complexity. We calculated a complexity index, in which a model received one point for each aspect that increased model complexity in comparison to a single-pathogen model. A) The frequency of complexity scores in our data set; black line represents the maximum possible score of 18. Mid-range scores between 4 and 7 were most common. B) The frequency of each complexity component in our data set. C) The co-occurrence of different complexity components within the same model. The inclusion of coinfected hosts and external interventions, in particular interventions, was very common in our data set. D) The correlation coefficients for each pair of complexity components. Question 23d was not answered for any models in our data set and thus was omitted from panels C and D.

Certain components of our complexity index were more likely to be included in the same model than others. Including co-infected hosts (question 23) and differing levels of susceptibility to other pathogens between uninfected and mono-infected hosts (question 24) were observed together more than any other pair of complexity score elements (51.5%, 160/311), as well as incorporating both co-infected hosts (question 23) and interventions (question 27) (48.9%, 152/311) and incorporating co-infected hosts and external factors impacting transmission (question 30) (48.9%, 152/311) (Figure 6C). When looking at the correlation of different components (either jointly present or absent in a model), the most highly correlated pairs were likely due to question redundancy and dependency in our survey. For example, questions 23a, “Does the model structure explicitly include within-host pathogen-pathogen interaction?” and 23b “What type of pathogen-pathogen interaction is represented?” are highly correlated (r=0.7). Other than these types of related questions, there were few very strong correlations, either positive or negative, indicating high variation in which aspects of co-circulation models in our data set elaborated on (Figure 6D).

## 4. Discussion

Pathogens do not exist in a vacuum; they are part of a complex ecosystem that, among other players, contains multiple pathogens that often vie for the same pool of susceptible hosts and can be influenced by the same external factors, such as climate and vector abundance. Mathematical models that explicitly explore the dynamics of co-circulating pathogens have the potential to inform numerous open questions in disease ecology and public health. In our systematic review of co-circulating pathogen models, we found that the scope of the literature in this field has gradually expanded over the past three decades. The emergence of HIV and the clear connection between HIV infection and increased susceptibility to other pathogens was a major catalyzing event for this area of work. However, even as models of multiple pathogens have become more common, there are still extensions of these models that remain underexplored, such as tying within-host processes to population-level models, exploring the effects of misdiagnosis of clinically similar diseases, and assessing the impacts of changes in resource allocation during outbreaks.

### 4.1 Model complexity

We created a complexity index to capture the extent to which a co-circulating pathogen model diverged from a simple, single-pathogen model, and found that the majority of models in our review fell into a middle range of this index. The inclusion of a second pathogen in the model often necessitated a few key additional parameters to capture the known biological interactions between the two pathogens, but a full range of model enhancements was rarely deployed. For example, including HIV and TB in the same model necessitates that the model structure account for susceptibility to TB in HIV-infected hosts. Other interactions between the two pathogens were also commonly included including higher likelihood of TB reactivation from the latent phase in HIV-infected individuals and increased morbidity and mortality in HIV-TB co-infecteds. Far less common in our data set was the inclusion of external factors that reflect the ways in which our health systems interface with the world of co-circulating pathogens and their hosts.

For example, arboviruses such as dengue, chikungunya, and Zika can present with similar clinical symptoms, which can lead to misdiagnosis of one infection for one another (Silva et al. 2019). Misdiagnoses can then result in spurious surveillance data from which we build our models (Oliveira et al. 2020), and potentially mis-appropriated public health funds targeted to prevent one disease at the expense of the others. Another topic that came hurtling to the foreground with the COVID-19 pandemic is the potential of one emerging pathogen to interrupt and delay healthcare services targeted towards the treatment and prevention of other pathogens (Bigouette et al. 2022). During the COVID-19 response, routine vaccination and surveillance programs for endemic diseases were put on hold in many nations, with negative consequences, especially for children (Menendez et al. 2020, Cardoso Pinto et al. 2022).

### 4.2 A new framework for model development

Model structure complexity often appeared to be driven by the authors’ scholarly motivation(s) for creating the model. Our analysis of author’s motivations resulted in binning papers into four meta-categories: (i) understanding the dynamics of co-circulating pathogens, (ii) developing novel mathematical models, (iii) implementing intervention or control measures, and (iv) calculating cost-effectiveness and optimal control of interventions. As an overarching schema to approach the current state of the field, we propose that papers in categories (i) and (ii) be classified under an umbrella of “**exploration**” and those in (iii) and (iv) as “**evaluation**.” This overarching classification provides a convenient framework to be used for both viewing the current state of the literature and evaluating where knowledge gaps remain. Many of the papers focused on **exploration** had a primarily mathematical goal to simply develop a compartmental model of two co-circulating pathogens (Lusiana et al. 2017, Jose et al. 2023), but some extended into testing more fundamental hypotheses, such as why two co-circulating pathogens can co- exist in some regions and not others (Amaku et al. 2011), or how asymmetric cross-immunity between related pathogens observed in laboratory studies may impact population-level transmission and prevalence (Restif et al. 2008). Notable examples of **evaluation** included exploring the tradeoff between decreased population-level transmission of tuberculosis with extended drug therapy regimens vs. the selective pressure for drug-resistant strains (Kunkel et al. 2016), and examining the impact that a lack of interventions due to health services disruptions from the COVID-19 pandemic had on diseases such as HIV, malaria, and tuberculosis (Hogan et al. 2020). Overall, this framework of exploration and evaluation can be useful for researchers to take stock of the current state of knowledge in their system and identify critical knowledge gaps to push the field forward.

### 4.3 Model quality

Despite the number of yet unexplored avenues in the field of co-circulating pathogens left to pursue, in our review we noted that there was a glut of papers with very similar features that perhaps represent a saturation of this corner of the field. Specifically, 40.8% (127/311) of papers included two pathogens and had near-identical results sections that calculated the disease-free equilibria in the model, the basic reproduction number, and performed one or two simple numerical simulations of host prevalence. These same models often fully lacked any data to inform the model (25.2%, 32/127), or used recycled parameters from the literature (72.4%, 92/127). Although there is value in such mathematical approaches, each additional study that performs these steps on a nearly identical system makes only a very incremental contribution to the body of knowledge of co-circulating pathogen dynamics.

Another quality issue we frequently noted was the misunderstanding and/or mischaracterizing of known biology in the construction of some models. Concrete examples included misalignment of stated model goals with reported model outcomes. For example, multiple papers stated in the introduction section that a goal of the model was to explore how HIV-related changes in host susceptibility to other pathogens may change population trends, but then nothing in the structure of the model reflected a change in host susceptibility. Similarly, authors would often expound on the importance of specific host cohorts in transmission, such as men who have sex with men or intravenous drug users, but then only explicitly include heterosexual sexual encounters in their contact process represented in the model. These overlooked errors in the pairing of model structure to known biology are worrisome in a peer-reviewed system. In addition to both authors and reviewers devoting more attention to detail, these errors suggest a need for greater emphasis on the correspondence between model structure and stated biological assumptions in training around mathematical modeling of infectious disease dynamics.

### 4.4 Future directions: Multi-scale, multi-pathogen models

Much of the complexity of disease dynamics arises from interactions across scales of biological organization; from within hosts to the population level, and from the population to the greater ecological community, disease dynamics occur across orders of magnitude. As the field of modeling co-circulating pathogens continues to grow and evolve, the effects of scale in modeling studies cannot be minimized. New studies are needed that examine multi-scale interactions and impacts of multiple pathogens in a single model. Consider, for example, the known effect of influenza on respiratory immune defenses, leading to a greater likelihood of infection by bacterial pathogens such as *Streptococcus pneumoniae* and also potentially worse clinical outcomes (Herrera et al. 2016). While a few papers in our data set did make simple adjustments to the model to include aspects such as changes in host susceptibility due to infection, very few contained a true within-host model where the processes at play within the host had the potential to influence population-level transmission in a nuanced or mechanistic manner. Work that connects the known within-host, cellular-level impacts of pathogen-pathogen interactions (McCullers 2006) to population-level prevalence and disease burden could give insight into better surveillance tactics and treatment schemes to mitigate the negative impacts of these pathogens. Key to this pursuit are appropriate data sources at multiple scales, from the individual to the population level, to verify how pathogens are interacting within hosts and tie these data to phenomena observed with co-circulating pathogen transmission at the population level. For respiratory pathogens, the COVID-19 pandemic and the accompanying “tripledemic” of COVID-19, influenza, and RSV has, at least temporarily, heightened testing and surveillance and expanded the use of excellent tools such as wastewater data (Boehm et al. 2023, Luo et al. 2023, Petros et al. 2024). We hope for a continued focus on gathering high-quality data for these and other pathogens, which will allow researchers to build robust, data-informed models that can directly inform public health efforts.

### 4.5 Study limitations

It is important to emphasize that we restricted our review to include only models that contained a mechanistic representation of co-circulating pathogens of humans. This criterion allowed for an in-depth focus on this subset of the literature, but we recognize that there are still many insights to be found from non-mechanistic statistical and geospatial approaches, models of theoretical pathogens, and work done in animal and plant systems. Work on theoretical pathogens can explore central mechanisms that drive the patterns seen in co-circulating pathogens in human populations, such as how co-infection may (or may not) drive changes in pathogen virulence (Brown et al. 2002). Experimental manipulations in non-human systems that would not be feasible or ethical with human subjects can also yield fundamental insights into the interactions and mechanisms at play in a multi-pathogen world, such as how variability in infection risk can be more strongly influenced by co-infecting pathogens than other mechanisms such as host exposure to parasites and host body condition (Telfer et al. 2010). These examples highlight the depth of investigation that is possible in other host-pathogen systems, and we want to emphasize that despite the focus of this systematic review, the power of non-human studies to inform and inspire work on human pathogens should not be discounted. A tandem review of co-circulating pathogen models that did not meet our inclusion criteria, such as non-mechanistic models and those focused on non-human pathogens, could help further inform and advance approaches across systems.

### 4.6 Conclusion

Co-circulating pathogen models can be a highly useful tool in the public health toolkit when it comes to preventing and mitigating infectious disease. From the papers in our review, we found clear examples of concrete recommendations that arise from considering co-circulating pathogens, ranging from actionable recommendations based on a realistic public health budget (Goyal and Murray 2017) to screening recommendations to meet WHO targets for a particular disease (Macgregor et al. 2020). We believe many more insights remain to be discovered from future work on co-circulating pathogens. What level of surveillance is needed to track and forecast a pathogen of interest when it co-circulates with other pathogens that cause similar clinical symptoms, such as arboviruses? What testing and treatment regimens can minimize the population-level effects of co-circulating respiratory pathogens that are known to have within-host interactions? How might antibacterial treatment for one pathogen influence the trajectory of antibiotic resistance not just in the concerned species, but other co-circulating bacteria? We hope that this systematic review serves as a launching point for others to identify and tackle the most exciting and pressing challenges presented by co-circulating pathogens, and in doing so mitigate the burden of infectious diseases.

## Data Availability

All data included in the study will be made publicly available upon publication at a journal and are available by request before publication.

## Acknowledgements

This work was supported by the NIH National Institute of General Medical Sciences R35 MIRA program to T.A.P. (grant no. R35GM143029).

Co-circulating pathogens of humans: A systematic review of mechanistic transmission models

## Supplemental: Co-circulating pathogens of humans: A 827 systematic review of mechanistic transmission models

**Figure S1.**
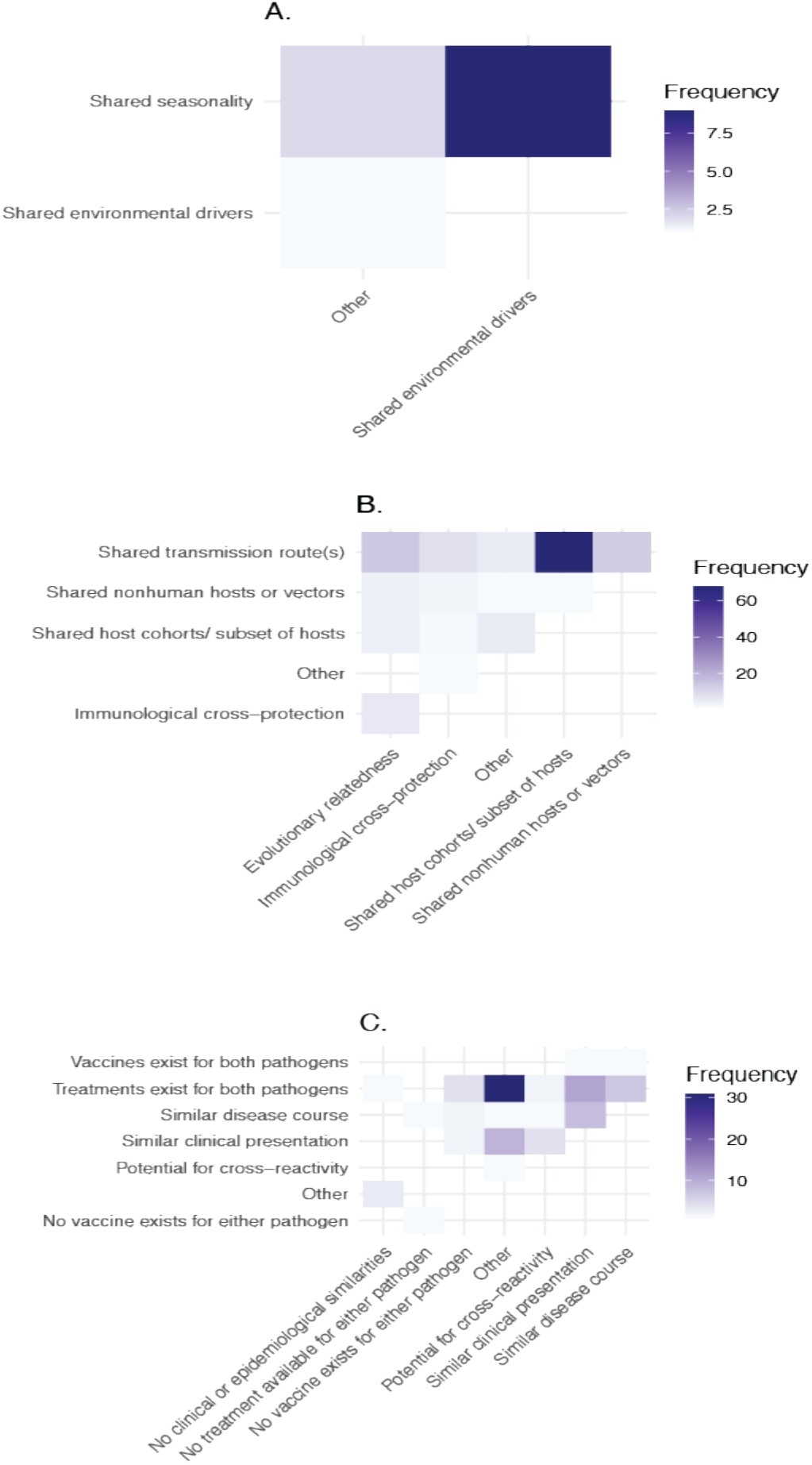
The co-occurrence of different environmental similarities (A), biological similarities (B), or clinical/epidemiological similarities (C) noted between pathogens in the same model

**Figure S2.**
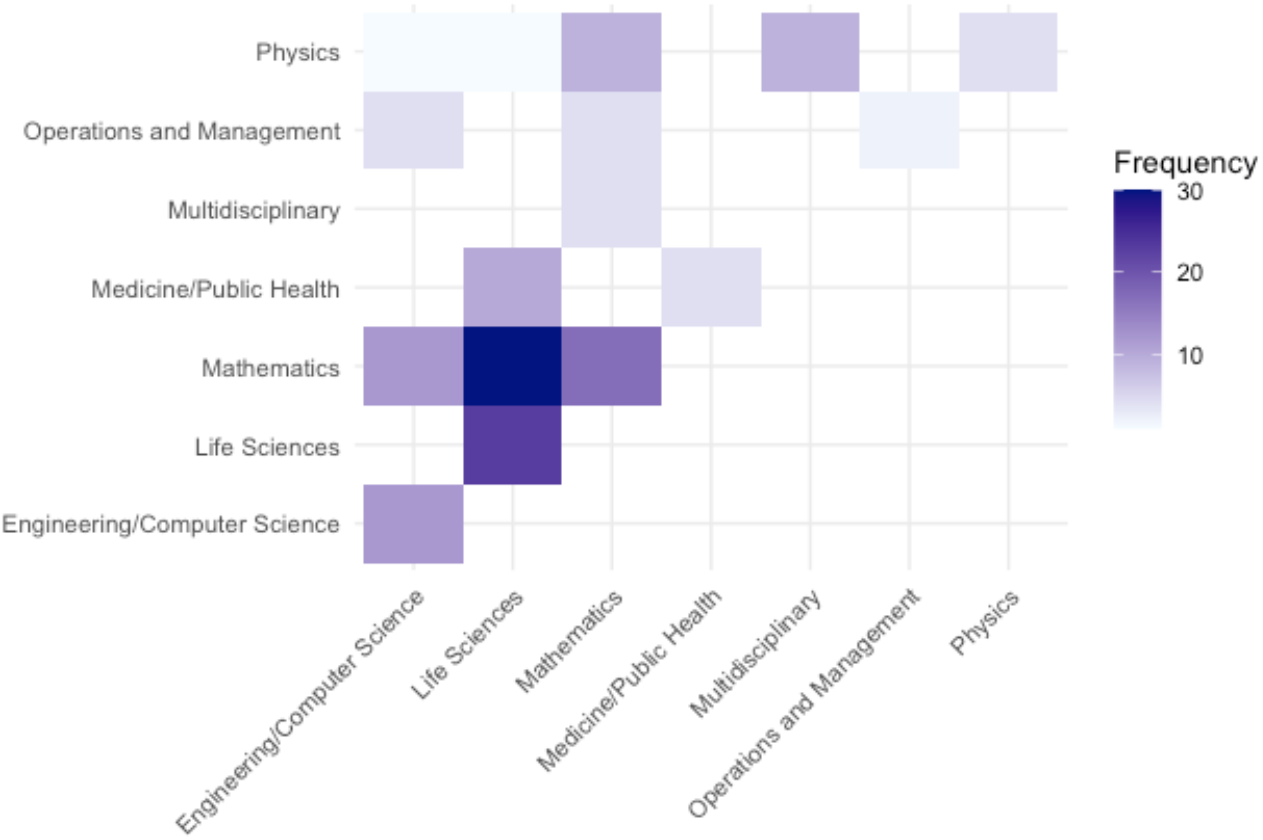
Overlapping classifications of journal types for the journals represented by papers in our data set.

**Table S1.**
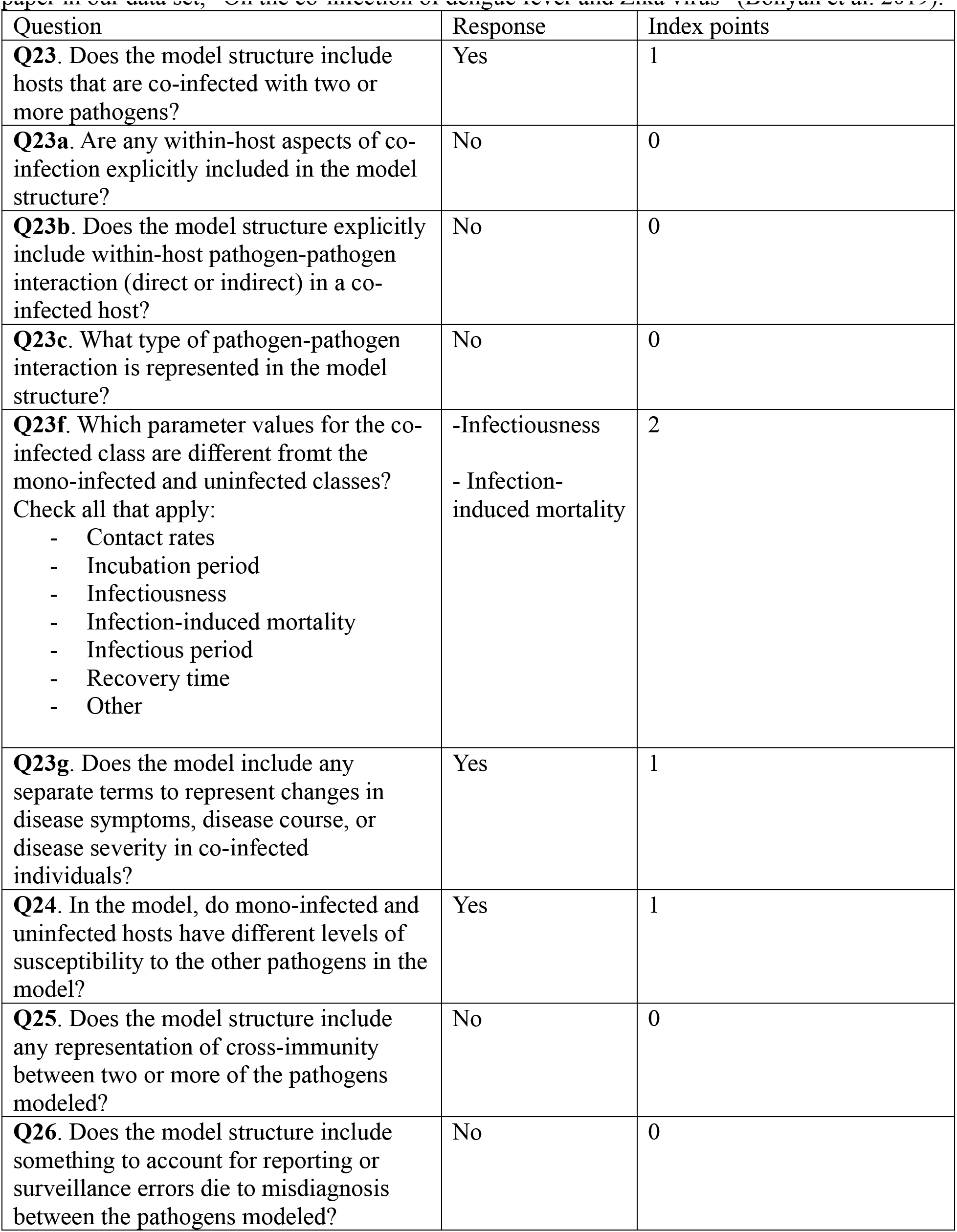
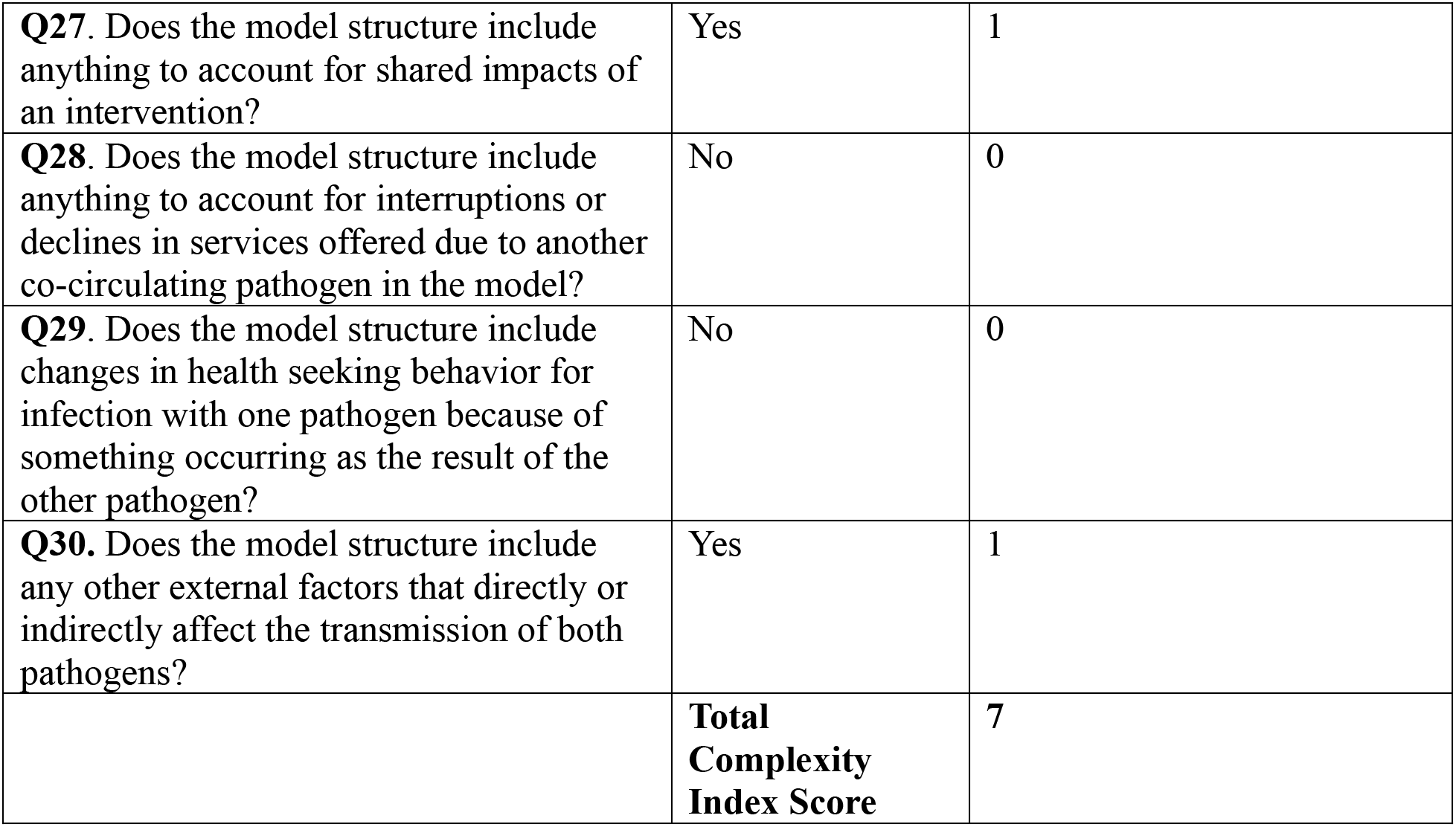
An example calculation of the complexity index. This calculation was made for one paper in our data set, “On the co-infection of dengue fever and Zika virus” (Bonyah et al. 2019).

| Question | Response | Index points |
| --- | --- | --- |
| **Q23.** Does the model structure include hosts that are co-infected with two or more pathogens? | Yes | 1 |
| **Q23a.** Are any within-host aspects of co-infection explicitly included in the model structure? | No | 0 |
| **Q23b.** Does the model structure explicitly include within-host pathogen-pathogen interaction (direct or indirect) in a co-infected host? | No | 0 |
| **Q23c.** What type of pathogen-pathogen interaction is represented in the model structure? | No | 0 |
| **Q23f.** Which parameter values for the co-infected class are different from the mono-infected and uninfected classes?<br>Check all that apply:<br>- Contact rates<br>- Incubation period<br>- Infectiousness<br>- Infection-induced mortality<br>- Infectious period<br>- Recovery time<br>- Other | -Infectiousness<br>- Infection-induced mortality | 2 |
| **Q23g.** Does the model include any separate terms to represent changes in disease symptoms, disease course, or disease severity in co-infected individuals? | Yes | 1 |
| **Q24.** In the model, do mono-infected and uninfected hosts have different levels of susceptibility to the other pathogens in the model? | Yes | 1 |
| **Q25.** Does the model structure include any representation of cross-immunity between two or more of the pathogens modeled? | No | 0 |
| **Q26.** Does the model structure include something to account for reporting or surveillance errors due to misdiagnosis between the pathogens modeled? | No | 0 |
| **Q27.** Does the model structure include anything to account for shared impacts of an intervention? | Yes | 1 |
| **Q28.** Does the model structure include anything to account for interruptions or declines in services offered due to another co-circulating pathogen in the model? | No | 0 |
| **Q29.** Does the model structure include changes in health seeking behavior for infection with one pathogen because of something occurring as the result of the other pathogen? | No | 0 |
| **Q30.** Does the model structure include any other external factors that directly or indirectly affect the transmission of both pathogens? | Yes | 1 |
|  | **Total Complexity Index Score** | **7** |

**Table S2.** We binned related Web of Science journal categories into meta categories used for analysis.s

| Web of Science Categories | Binned category |
| --- | --- |
| Mathematical and Computational Biology; Mathematics, Applied; Mathematics, Interdisciplinary Applications; Mathematics; Statistics and Probability | Mathematics |
| Computer Science, Interdisciplinary Applications; Computer Science, Software Engineering; Computer Science, Information Systems; Engineering, Electrical and Electronic; Engineering, Chemical; Engineering, Mechanical; Engineering, Multidisciplinary; Automation and Control Systems; Mechanics | Engineering/Computer Science |
| Management; Operations Research and Management Science | Operations and Management |
| Infectious Diseases; Biology; Ecology; Microbiology; Immunology; Parasitology; Tropical Medicine; Virology; Cell Biology; Genetics and Heredity; Environmental Science; Evolutionary Biology; Environmental Sciences; Biophysics; Biotechnology and Applied Microbiology; Biochemical Research Methods | Life Sciences |
| Gastroenterology and Hepatology; Medicine, General and Internal; Medicine, Research and Experimental; Medicine/Public Health; Substance Abuse; Psychiatry; Public, Environmental and Occupational Health | Medicine/Public Health |
| Physics, Applied; Physics, Mathematical; Physics, Multidisciplinary | Physics |
| Multidisciplinary Sciences; Materials Science, Multidisciplinary | Multidisciplinary |

## References

1. Abu-Raddad, L. J., L. Sabatelli, J. T. Achterberg, J. D. Sugimoto, I. M. Longini, C. Dye, and M. E. Halloran. 2009. Epidemiological benefits of more-eOective tuberculosis vaccines, drugs, and diagnostics. Proceedings of the National Academy of Sciences 106:13980–13985.

2. Alemu, A., Y. Shiferaw, Z. Addis, B. Mathewos, and W. Birhan. 2013. EOect of malaria on HIV/AIDS transmission and progression. Parasites & Vectors 6:18.

3. Amaku, M., F. A. B. Coutinho, and E. Massad. 2011. Why dengue and yellow fever coexist in some areas of the world and not in others? Biosystems 106:111–120.

4. Aria, M., T. Le, C. Cuccurullo, A. Belfiore, and J. Choe. 2024. openalexR: An R-Tool for Collecting Bibliometric Data from OpenAlex. The R Journal 15:167–180.

5. Bigouette, J. P., A. W. Callaghan, M. Donadel, A. M. Porter, L. Rosencrans, J. S. Lickness, S. Blough, X. Li, R. T. Perry, A. J. Williams, H. M. Scobie, B. A. Dahl, J. McFarland, and C. S. Murrill. 2022. EOects of COVID-19 on Vaccine-Preventable Disease Surveillance Systems in the World Health Organization African Region, 2020. Emerging Infectious Diseases 28.

6. Boehm, A. B., M. K. Wolfe, B. J. White, B. Hughes, D. Duong, and A. Bidwell. 2023. More than a Tripledemic: Influenza A Virus, Respiratory Syncytial Virus, SARS-CoV-2, and Human Metapneumovirus in Wastewater during Winter 2022–2023. Environmental Science & Technology Letters 10:622–627.

7. Bonyah, E., M. A. Khan, K. O. Okosun, and J. F. Gómez-Aguilar. 2019. On the co-infection of dengue fever and Zika virus. Optimal Control Applications and Methods 40:394–421.

8. Brooks, M. E., K. Kristensen, K. J. Van Benthem, A. Magnusson, C. W. Berg, A. Nielsen, H. J. Skaug, M. Machler, and B. M. Bolker. 2017. glmmTMB balances speed and flexibility among packages for zero-inflated generalized linear mixed modeling. The R journal 9:378–400.

9. Brown, S. P., M. E. Hochberg, and B. T. Grenfell. 2002. Does multiple infection select for raised virulence? Trends in Microbiology 10:401–405.

10. Cardoso Pinto, A. M., L. Ranasinghe, P. J. Dodd, S. S. Budhathoki, J. A. Seddon, and E. Whittaker. 2022. Disruptions to routine childhood vaccinations in low- and middle- income countries during the COVID-19 pandemic: A systematic review. Frontiers in Pediatrics 10:979769. ChatGPT. 2024, April 1. . OpenAI.

11. Cowger, T. L., E. J. Murray, J. Clarke, M. T. Bassett, B. O. Ojikutu, S. M. Sánchez, N. Linos, and K. T. Hall. 2022. Lifting Universal Masking in Schools — Covid-19 Incidence among Students and StaO. New England Journal of Medicine 387:1935–1946.

12. Da Silva Pessoa Vieira, C. J., C. Steiner São Bernardo, D. J. Ferreira Da Silva, J. Rigotti Kubiszeski, E. Serpa Barreto, H. A. De Oliveira Monteiro, G. R. Canale, C. A. Peres, A. L. Massey, T. Levi, and R. Vieira De Morais Bronzoni. 2022. Land-use eOects on mosquito biodiversity and potential arbovirus emergence in the Southern Amazon, Brazil. Transboundary and Emerging Diseases 69:1770–1781.

13. Eikenberry, S. E., and A. B. Gumel. 2018. Mathematical modeling of climate change and malaria transmission dynamics: a historical review. Journal of Mathematical Biology 77:857–933.

14. Ferguson, N. M., D. A. T. Cummings, C. Fraser, J. C. Cajka, P. C. Cooley, and D. S. Burke. 2006. Strategies for mitigating an influenza pandemic. Nature 442:448–452.

15. Galvin, S. R., and M. S. Cohen. 2004. The role of sexually transmitted diseases in HIV transmission. Nature Reviews Microbiology 2:33–42.

16. Gaythorpe, K. A., K. Abbas, J. Huber, A. Karachaliou, N. Thakkar, K. WoodruO, X. Li, S. Echeverria-Londono, VIMC Working Group on COVID-19 Impact on Vaccine Preventable Disease, A. Arsene Bita Fouda, F. Cutts, E. Dansereau, A. Durupt, U. GriOiths, J. Horton, L. K. Krause, K. Kretsinger, T. Mengistu, I. Mirza, S. R. Procter, S. Shendale, M. Ferrari, M. L. Jackson, K. McCarthy, T. A. Perkins, C. Trotter, and M. Jit. 2021. Impact of COVID-19-related disruptions to measles, meningococcal A, and yellow fever vaccination in 10 countries. eLife 10:e67023.

17. Goyal, A., and J. M. Murray. 2017. Roadmap to control HBV and HDV epidemics in China. Journal of Theoretical Biology 423:41–52.

18. Heesterbeek, H., R. M. Anderson, V. Andreasen, S. Bansal, D. De Angelis, C. Dye, K. T. D. Eames, W. J. Edmunds, S. D. W. Frost, S. Funk, T. D. Hollingsworth, T. House, V. Isham, P. Klepac, J. Lessler, J. O. Lloyd-Smith, C. J. E. Metcalf, D. Mollison, L. Pellis, J. R. C. Pulliam, M. G. Roberts, C. Viboud, and Isaac Newton Institute IDD Collaboration. 2015. Modeling infectious disease dynamics in the complex landscape of global health. Science 347:aaa4339.

19. Herrera, A. L., V. C. Huber, and M. S. Chaussee. 2016. The Association between Invasive Group A Streptococcal Diseases and Viral Respiratory Tract Infections. Frontiers in Microbiology 7.

20. Hogan, A. B., B. L. Jewell, E. Sherrard-Smith, J. F. Vesga, O. J. Watson, C. Whittaker, A. Hamlet, J. A. Smith, P. Winskill, R. Verity, M. Baguelin, J. A. Lees, L. K. Whittles, K. E. C. Ainslie, S. Bhatt, A. Boonyasiri, N. F. Brazeau, L. Cattarino, L. V. Cooper, H. Coupland, G. Cuomo-Dannenburg, A. Dighe, B. A. Djaafara, C. A. Donnelly, J. W. Eaton, S. L. Van Elsland, R. G. FitzJohn, H. Fu, K. A. M. Gaythorpe, W. Green, D. J. Haw, S. Hayes, W. Hinsley, N. Imai, D. J. Laydon, T. D. Mangal, T. A. Mellan, S. Mishra, G. Nedjati-Gilani, K. V. Parag, H. A. Thompson, H. J. T. Unwin, M. A. C. Vollmer, C. E. Walters, H. Wang, Y. Wang, X. Xi, N. M. Ferguson, L. C. Okell, T. S. Churcher, N. Arinaminpathy, A. C. Ghani, P. G. T. Walker, and T. B. Hallett. 2020. Potential impact of the COVID-19 pandemic on HIV, tuberculosis, and malaria in low-income and middle-income countries: a modelling study. The Lancet Global Health 8:e1132– e1141.

21. Huang, Y., and P. Rohani. 2006. Age-structured eOects and disease interference in childhood infections. Proceedings of the Royal Society B: Biological Sciences 273:1229–1237.

22. Isea, R., and K. Lonngren. 2016. A Preliminary Mathematical Model for the Dynamic Transmission of Dengue, Chikungunya and Zika. American Journal of Modern Physics and Application 3:11–15.

23. Jenness, S. M., K. M. Weiss, S. M. Goodreau, T. Gift, H. Chesson, K. W. Hoover, D. K. Smith, A. Y. Liu, P. S. Sullivan, and E. S. Rosenberg. 2017. Incidence of Gonorrhea and Chlamydia Following Human Immunodeficiency Virus Preexposure Prophylaxis Among Men Who Have Sex With Men: A Modeling Study. Clinical Infectious Diseases 65:712–718.

24. Johnson, L. F., R. E. Dorrington, D. Bradshaw, and D. J. Coetzee. 2011. The eOect of syndromic management interventions on the prevalence of sexually transmitted infections in South Africa. Sexual & Reproductive Healthcare 2:13–20.

25. Jose, S. A., R. Raja, B. I. Omede, R. P. Agarwal, J. Alzabut, J. Cao, and V. E. Balas. 2023. Mathematical modeling on co-infection: transmission dynamics of Zika virus and Dengue fever. Nonlinear Dynamics 111:4879–4914.

26. Kate K Orroth, Esther E Freeman, Roel Bakker, Anne Buvé, Judith R Glynn, Marie-Claude Boily, Richard G White, J Dik F Habbema, and Richard J Hayes. 2007. Understanding the diOerences between contrasting HIV epidemics in east and west Africa: results from a simulation model of the Four Cities Study. Sexually Transmitted Infections 83:i5.

27. Korenromp, E. L., R. G. White, K. K. Orroth, R. Bakker, A. Kamali, D. Serwadda, R. H. Gray, H. Grosskurth, J. D. F. Habbema, and R. J. Hayes. 2005. Determinants of the Impact of Sexually Transmitted Infection Treatment on Prevention of HIV Infection: A Synthesis of Evidence from the Mwanza, Rakai, and Masaka Intervention Trials. The Journal of Infectious Diseases 191:S168–S178.

28. Kunkel, A., F. W. Crawford, J. Shepherd, and T. Cohen. 2016. Benefits of continuous isoniazid preventive therapy may outweigh resistance risks in a declining tuberculosis/HIV coepidemic. AIDS 30:2715–2723.

29. LaOerty, K. D. 2009. The ecology of climate change and infectious diseases. Ecology 90:888–900.

30. Lemaitre, J., D. Pasetto, J. Perez-Saez, C. Sciarra, J. F. Wamala, and A. Rinaldo. 2019. Rainfall as a driver of epidemic cholera: Comparative model assessments of the eOect of intra-seasonal precipitation events. Acta Tropica 190:235–243.

31. Lloyd-Smith, J. O., M. Poss, and B. T. Grenfell. 2008. HIV-1/parasite co-infection and the emergence of new parasite strains. Parasitology 135:795–806.

32. Luo, W., Q. Liu, Y. Zhou, Y. Ran, Z. Liu, W. Hou, S. Pei, and S. Lai. 2023. Spatiotemporal variations of “triple-demic” outbreaks of respiratory infections in the United States in the post-COVID-19 era. BMC Public Health 23:2452.

33. Lusiana, V., P. S. Putra, N. Nuraini, and E. Soewono. 2017. Mathematical modeling of transmission co-infection tuberculosis in HIV community. Page 020012. Makassar, Indonesia.

34. Macgregor, L., M. Desai, N. K. Martin, J. Nicholls, F. Hickson, P. Weatherburn, M. Hickman, and P. Vickerman. 2020. Scaling up screening and treatment for elimination of hepatitis C among men who have sex with men in the era of HIV pre-exposure prophylaxis. EClinicalMedicine 19:100217.

35. Marimuthu, Y., B. Nagappa, N. Sharma, S. Basu, and K. K. Chopra. 2020. COVID-19 and tuberculosis: A mathematical model based forecasting in Delhi, India. Indian Journal of Tuberculosis 67:177–181.

36. Mayer, K. H., C. L. Karp, P. G. Auwaerter, and K. H. Mayer. 2007. Coinfection with HIV and Tropical Infectious Diseases. I. Protozoal Pathogens. Clinical Infectious Diseases 45:1208–1213.

37. McCullers, J. A. 2006. Insights into the Interaction between Influenza Virus and Pneumococcus. Clinical Microbiology Reviews 19:571–582.

38. Mendenhall, E., B. A. Kohrt, C. H. Logie, and A. C. Tsai. 2022. Syndemics and clinical science. Nature Medicine 28:1359–1362.

39. Menendez, C., R. Gonzalez, F. Donnay, and R. G. F. Leke. 2020. Avoiding indirect eOects of COVID-19 on maternal and child health. The Lancet Global Health 8:e863–e864.

40. Montales, M. T., A. Chaudhury, A. Beebe, S. Patil, and N. Patil. 2015. HIV-Associated TB Syndemic: A Growing Clinical Challenge Worldwide. Frontiers in Public Health 3.

41. Mordecai, E. A., J. M. Caldwell, M. K. Grossman, C. A. Lippi, L. R. Johnson, M. Neira, J. R. Rohr, S. J. Ryan, V. Savage, M. S. Shocket, R. Sippy, A. M. Stewart Ibarra, M. B. Thomas, and O. Villena. 2019. Thermal biology of mosquito-borne disease. Ecology Letters 22:1690–1708.

42. Oidtman, R. J., G. España, and T. A. Perkins. 2021. Co-circulation and misdiagnosis led to underestimation of the 2015–2017 Zika epidemic in the Americas. PLOS Neglected Tropical Diseases 15:e0009208.

43. Okuneye, K. O., J. X. Velasco-Hernandez, and A. B. Gumel. 2017. The “Unholy” Chikunginya-Dengue-Zika Trinity: A Theoretical Analysis. Journal of Biological Systems 25:545–585.

44. Oliveira, J. F., M. S. Rodrigues, L. M. Skalinski, A. E. S. Santos, L. C. Costa, L. L. Cardim, E. S. Paixão, M. D. C. N. Costa, W. K. Oliveira, M. L. Barreto, M. G. Teixeira, and R. F. S. Andrade. 2020. Interdependence between confirmed and discarded cases of dengue, chikungunya and Zika viruses in Brazil: A multivariate time-series analysis. PLOS ONE 15:e0228347.

45. Pagel, C., and C. A. Yates. 2022. Role of mathematical modelling in future pandemic response policy. BMJ:e070615.

46. Petros, B. A., C. E. Milliren, P. C. Sabeti, and A. OzonoO. 2024. Increased Pediatric Respiratory Syncytial Virus Case Counts Following the Emergence of Severe Acute Respiratory Syndrome Coronavirus 2 Can Be Attributed to Changes in Testing. Clinical Infectious Diseases:ciae140.

47. R Core Team. 2021. R: A language and environment for statistical computing. R Foundation for Statistical Computing, Vienna, Austria.

48. Rao, D. W., C. J. Bayer, G. Liu, A. Chikandiwa, M. Sharma, C. L. Hathaway, N. Tan, N. Mugo, and R. V. Barnabas. 2022. Modelling cervical cancer elimination using single-visit screening and treatment strategies in the context of high HIV prevalence: estimates for KwaZulu-Natal, South Africa. Journal of the International AIDS Society 25:e26021.

49. Restif, O., D. N. Wolfe, E. M. Goebel, O. N. Bjornstad, and E. T. Harvill. 2008. Of mice and men: asymmetric interactions between Bordetella pathogen species. Parasitology 135:1517–1529.

50. Ribas Freitas, A. R., A. A. Pinheiro Chagas, A. M. Siqueira, and L. Pamplona De Góes Cavalcanti. 2024. How much of the current serious arbovirus epidemic in Brazil is dengue and how much is chikungunya? The Lancet Regional Health - Americas 34:100753. Richard Orr, Heather Piwowar, Jason Priem. 2024. Unpaywall. OurResearch.

51. Rohani, P., C. J. Green, N. B. Mantilla-Beniers, and B. T. Grenfell. 2003. Ecological interference between fatal diseases. Nature 422:885–888.

52. Silva, M. M. O., L. B. Tauro, M. Kikuti, R. O. Anjos, V. C. Santos, T. S. F. Gonçalves, I. A. D. Paploski, P. S. S. Moreira, L. C. J. Nascimento, G. S. Campos, A. I. Ko, S. C. Weaver, M. G. Reis, U. Kitron, and G. S. Ribeiro. 2019. Concomitant Transmission of Dengue, Chikungunya, and Zika Viruses in Brazil: Clinical and Epidemiological Findings From Surveillance for Acute Febrile Illness. Clinical Infectious Diseases 69:1353–1359.

53. Telfer, S., X. Lambin, R. Birtles, P. Beldomenico, S. Burthe, S. Paterson, and M. Begon. 2010. Species Interactions in a Parasite Community Drive Infection Risk in a Wildlife Population. Science 330:243–246.

54. Thomson, M. C., F. J. Doblas-Reyes, S. J. Mason, R. Hagedorn, S. J. Connor, T. Phindela, A. P. Morse, and T. N. Palmer. 2006. Malaria early warnings based on seasonal climate forecasts from multi-model ensembles. Nature 439:576–579.

55. Tsai, A. C., E. Mendenhall, J. A. Trostle, and I. Kawachi. 2017. Co-occurring epidemics, syndemics, and population health. The Lancet 389:978–982.

56. Vickerman, P., N. K. Martin, and M. Hickman. 2012. Understanding the trends in HIV and hepatitis C prevalence amongst injecting drug users in diOerent settings— Implications for intervention impact. Drug and Alcohol Dependence 123:122–131.

57. Vitoria, M., R. Granich, C. F. Gilks, C. Gunneberg, M. Hosseini, W. Were, M. Raviglione, and K. M. De Cock. 2009. The Global Fight Against HIV/AIDS, Tuberculosis, and Malaria. American Journal of Clinical Pathology 131:844–848.

58. de Vos, A. S., J. J. van der Helm, M. Prins, and M. E. Kretzschmar. 2012. Determinants of persistent spread of HIV in HCV-infected populations of injecting drug users. Epidemics 4:57–67. Web of Science API. 2024. . Clarivate.

59. Zahouli, J. B. Z., B. G. Koudou, P. Müller, D. Malone, Y. Tano, and J. Utzinger. 2017. EOect of land-use changes on the abundance, distribution, and host-seeking behavior of Aedes arbovirus vectors in oil palm-dominated landscapes, southeastern Côte d’Ivoire. PLOS ONE 12:e0189082.

60. Zelenev, A., J. Li, A. Mazhnaya, S. Basu, and F. L. Altice. 2018. Hepatitis C virus treatment as prevention in an extended network of people who inject drugs in the USA: a modelling study. The Lancet Infectious Diseases 18:215–224.

